# Intracerebral Hemorrhage Outcomes after Reversal of Subtherapeutic Warfarin: Analysis of Data from GWTG-Stroke

**DOI:** 10.1101/2024.12.18.24319293

**Authors:** Aaron N. LacKamp, Jeremy M. Weber, Brian C. Mac Grory, Adrien J. Caye, Chaeli Stenuf, Eric E. Smith, Justin D. Daniels, Tiffany T Barkley, Steven R. Messe, Brooke Alhanti, Kevin N. Sheth, Rosalia G Blanco, Gregg C. Fonarow, Ying Xian, Halinder S. Mangat

## Abstract

**Background:** Current guidelines recommend reversal of warfarin anticoagulation with intracranial hemorrhages. The benefit of reversing subtherapeutic warfarin anticoagulation in acute spontaneous intracerebral hemorrhage (ICH) is uncertain.

**Methods:** An observational cohort used Get With The Guidelines® Stroke registry between January 2015 and January 2022 to determine association of reversal with outcomes for subtherapeutic anticoagulation (INR 1.5 to 1.9). Exclusions were thrombolytics, direct oral anticoagulants, transferring out, or leaving against medical advice.

**Results:** The primary outcome (mRS 0-3 at discharge) was assessed among 1868 patients (mean age 73 years, 42% female), which occurred in 188/894 (21.0%) with reversal and 225/974 (23.1%) without reversal (adjusted odds ratio (aOR) 0.80 [95% CI 0.63-1.005]). Ordinal analysis showed higher odds of mRS 0-4 vs. 5-6 with reversal {52.7% vs 42.5% (aOR 1.21 [1.001, 1.48])}. Outcomes not requiring mRS were analyzed among 2569 patients. Mortality or discharge to hospice was lower with reversal {30.6% vs 41.5% (aOR 0.75 [95% CI, 0.63, 0.89])}. Fewer were ambulatory at discharge {25.8% vs 35.7% (aOR 0.68 [0.54, 0.85])}, fewer discharged to home {18.4% vs 21.7% (aOR 0.79 [0.65, 0.97])}, more discharged to skilled nursing {21.0% vs 15.7% (aOR 1.33 [1.08, 1.65])}, and more discharged to rehabilitation {24.9% vs 18.9% (aOR 1.20 [0.98, 1.47])}. Hospital length of stay was longer {median 6 vs 4 days (adjusted risk ratio (aRR) 1.25 [95% CI, 1.13, 1.37]). There was no difference in venous thromboembolism {2.9% vs 2.3% (aOR 1.47 [0.88, 2.46])}.

**Discussion:** Reversal of subtherapeutic warfarin with acute spontaneous intracerebral hemorrhage and INR 1.5 - 1.9 was not associated with improvement in functional outcome based upon discharge mRS 0-3 vs 4-6. Patients that received a reversal agent had 25% lower odds of dying in the hospital or being discharged to hospice, but had a longer hospital stay and were less likely to be fully ambulatory at discharge.

## Introduction

Intracerebral hemorrhage is a highly morbid condition. The primary neurologic injury occurs at the time of the bleeding event. Patients with subsequent neurologic deterioration have worse long-term outcomes.^1,2^ Hematoma expansion is a major cause of neurologic deterioration within the first 24 hours^3,4^ followed by cerebral edema formation in the first 72 hours,^1^ both are related to the total intracranial hematoma volume. Two interventions are widely used to limit hematoma expansion and the subsequent total bur^-^ den of disease^4,5^ and disability:^6^ correction of any coagulopathy,^7,8,9^ and reduction of elevated blood pressure.^6,10,11,12,13,14^

Recent guidelines are available for intracranial hemorrhage and recommend reversal of vitamin K antagonists when patients are fully anticoagulated with INR >2.0.^15,16,17^ Intracerebral hemorrhage constitutes a large proportion of these patients. However some patients present with less than full anticoagulation (INR 1.5-1.9). In this population the benefit or harm of administering reversal agents has not been investigated. The expectation of benefit is not obvious for several reasons: the magnitude of the effect of the primary injury may outweigh any benefit of reversal, the effect of reversal on INR may diminish when values are closer to normal, and there may be increased thrombotic complications or volume overload when administering agents to reverse anticoagulation. For example, the reversal of antiplatelet agents in the PATCH trial resulted in increased mortality and disability although the reasons for this finding are unclear.^18^ Lastly, vitamin K dependent factor (II, VII, IX, X) activities do not decrease below 50% of normal activity until INR is greater than 1.5.^19^

There is clinical equipoise whether to provide reversal for subtherapeutic warfarin anticoagulation in patients with intracerebral hemorrhage. The aim of this investigation was to determine whether reversal is associated with functional outcome, mortality, or thrombotic events in patients taking warfarin who have intracerebral hemorrhage and incomplete anticoagulation (INR1.5 to 1.9).

## Methods

### Study Design

This observational cohort study analyzes data from the Get With The Guidelines® Stroke (GWTG-Stroke) registry^20^ between January 1, 2015 and January 4, 2022. Statistical analysis was performed in March through May 2022.

The Duke Clinical Research Institute served as the data analysis center and has an agreement to analyze the aggregate de-identified data for research purposes. The Institutional Review Board at Duke University Health approved this study. Each participating hospital received either human research approval to enroll cases without individual patient consent under the common rule, or a waiver of authorization and exemption from subsequent review by their institutional review board. We followed the Strengthening the Reporting of Observational Studies in Epidemiology (STROBE)^21^ reporting guidelines as a guide.

### Study Population

#### Inclusion criteria

Patients were included if admitted on or after January 1, 2015 through January 4, 2022 for an intracerebral hemorrhage to a hospital reporting on the comprehensive stroke center form. Intracerebral hemorrhage is one of six discrete final diagnoses for cases included in the GWTG-Stroke ® registry. Cases due to trauma and underlying vascular malformations are excluded by registry guidelines. The reporting hospitals must be fully participating (hospital-level medical history completion rate > 75%). Patients must have been on warfarin with INR 1.5 to 1.9 at admission. INR was rounded to two significant figures before applying the inclusion criterion.

#### Exclusion criteria

The following exclusions were applied: direct oral anticoagulant (DOAC) prior to admission, patients transferred out of the reporting hospital, patients who left against medical advice, patients for whom thrombolytic was given, and patients with missing data for gender, INR, mRS at discharge, or warfarin use. The absence of a recorded gender was used to screen for incomplete records and is consistent with other GWTG-Stroke analyses. There were no specific analyses based upon sex or gender. Please refer to Figure 1 for an explanation of how the patient cohorts were determined.

**Figure 1.**
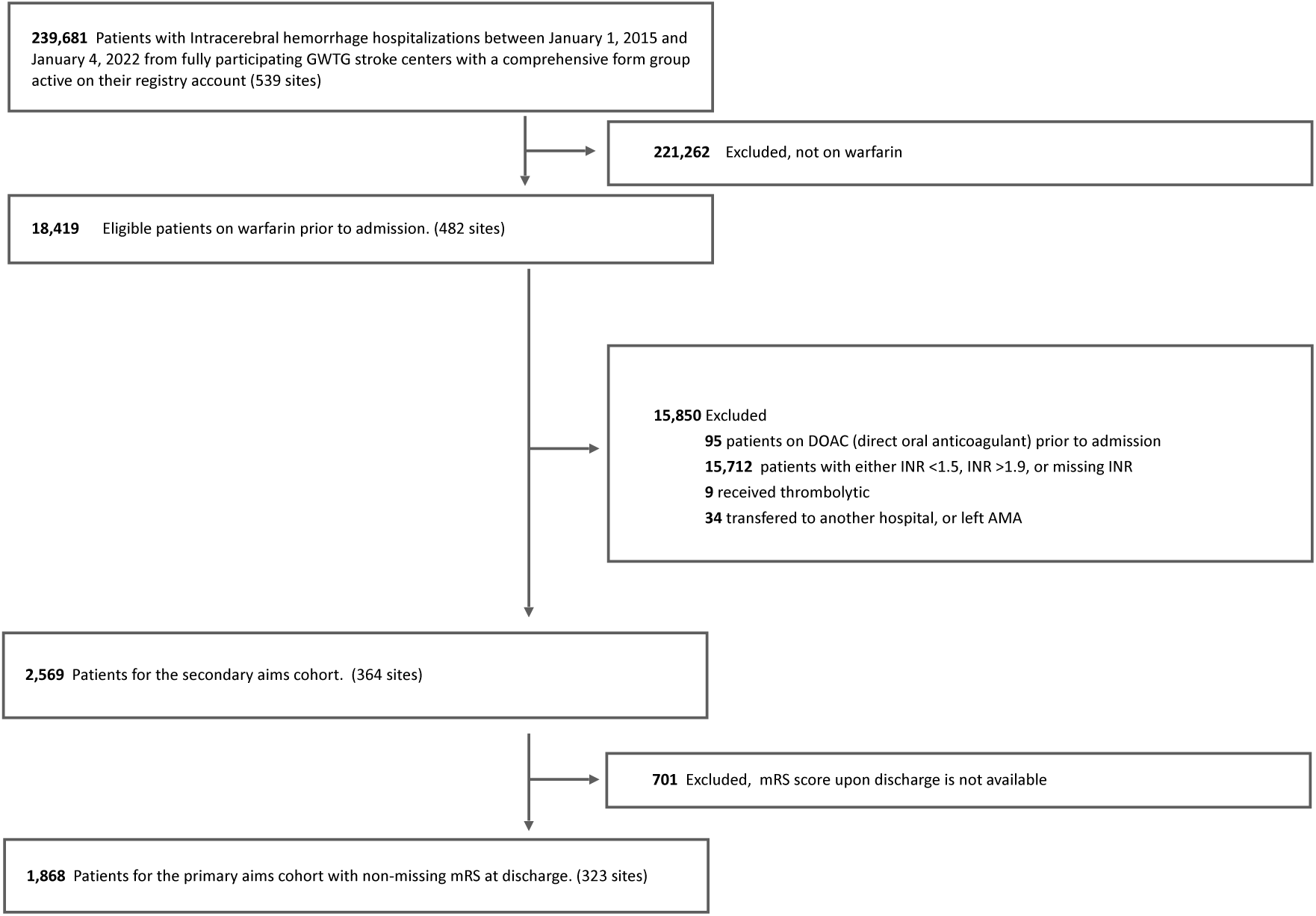
Study cohort flow chart. This figure is a visual representation of study participants from the Get With the Guidelines - Stroke (GWTG-Stroke) study population and includes the breakdown of how the study population was refined based upon predetermined exclusion criteria. AMA indicates against medical advice.

### Primary outcome

Dichotomized Modified Rankin Score (mRS) at discharge was used as the primary outcome. The mRS is an ordinal score of disability and dependence after stroke on a scale of 0 (no effect) to 6 (death). Dichotomization of mRS is sometimes performed at 0-3 vs. 4-6 for studies in highly morbid neurologic conditions such as intracerebral hemorrhage or severe stroke.^22,23,24^

### Secondary outcomes

In-hospital mortality was combined with discharge to hospice as a secondary outcome. Patients with documented comfort measures only were counted as an event for this composite of in-hospital mortality or discharge to hospice outcome. Other secondary aims included discharge mRS on an ordinal scale (0, 1, 2, 3, 4, 5-6), deep vein thrombosis (DVT) or pulmonary embolism (PE), hospital length of stay (LOS), hospital LOS of those that survived to discharge, discharge disposition (home, hospice/death, skilled nursing, rehab), ambulation status at discharge (fully ambulatory vs. all other states). All measures were counted at the time of discharge. To determine any influence of disease severity, we tested for interaction between ICH score (0-1, 2-3, 4-6, and unknown) and the effect of reversal agents on discharge mRS as both dichotomized (0-3 vs 4-6) and ordinal values (0, 1, 2, 3, 4, 5-6), the composite outcome of in-hospital mortality or discharge to hospice, and ambulatory status at discharge.

### Handling of missing data

Patients were excluded if missing data for INR, mRS at discharge, or warfarin usage. For medical history variables missing values were interpreted as not present. Hospital characteristics were not imputed. Model covariates with large proportions of missing values (25% or more) were excluded from the analysis. For variables with fewer than 25% missing values multiple imputation was used with the Full Conditional Specification method. Purely descriptive data not used in models were not imputed. Missing values for anticoagulant reversal were assumed to mean no reversal was given. Missing values in documented DVT/PE was assumed to mean that the complication did not occur.

### Statistical Analysis

Patient and hospital characteristics were described by whether a reversal agent was administered during hospitalization. Continuous variables were summarized using mean (SD), median (25^th^ percentile, 75^th^ percentile), and range. Categorical variables are presented as n/N (percentage). All percentages were calculated out of the number of patients with non-missing data for that variable.

To derive propensity scores, a multivariable logistic regression model was fit with reversal of anticoagulation as the dependent variable and included the following covariates: age, gender, race/ethnicity, hypertension, diabetes, heart failure, CAD/MI, prior ischemic stroke, renal insufficiency, atrial fibrillation, smoking, alcohol/drug abuse, sleep apnea, antiplatelet medications, antihypertensive medications, arrival via EMS, arrival during off-hours, admission GCS, ICH score, NIHSS, BMI, weakness/paresis, altered LOC, aphasia, systolic BP, diastolic BP, heart rate, INR, creatinine, hospital region, annual ICH stroke volume, and Joint Commission status as primary or comprehensive stroke center.

Subjects were weighted in proportion to the probability that they belonged to the opposite group (overlap weighting) for all models. The propensity score model for the primary aim and mRS-related secondary aims was modeled using the 1868 patients with nonmissing mRS. The propensity score model for the non-mRS secondary aims used 2569 patients (patients with and without discharge mRS available). Both propensity score models used the same list of covariates. For the primary aim, we determined the association between reversal of anticoagulation and discharge mRS of 0-3 using logistic regression. When considering mRS as an ordinal variable the proportional odds assumption was violated therefore a non-proportional odds cumulative logistic regression model was fit. The model generated five odds ratios comparing better vs. worse mRS (e.g. mRS 0 vs. 1-6, 0-1 vs. 2-6,…0-4 vs. 5-6) and allowed assessment of the impact of INR reversal across the spectrum of categories. Logistic regression was used to model the composite of in-hospital mortality or discharge to hospice, development of DVT or PE, the four binary variables for discharge disposition, and ambulation status at discharge.

Negative binomial regression model was fit for the LOS. All statistical analyses were performed in SAS 9.4 and figures were made in R 4.1.3.

### Sensitivity analysis for the primary aim

As a sensitivity analysis, we refit the propensity model using INR <= 1.4 after reversal in place of reversal itself. A similar logistic regression model as the primary aim was refit using these new weights and changing the independent variable.

### Interaction with ICH Score

We tested an interaction between reversal and ICH score (0-1, 2-3, 4-6, and unknown) to determine the effect on mRS at discharge, combined in-hospital-mortality-and-discharge-to-hospice, and ambulatory status at discharge varied by disease severity.

## Results

Patient and hospital characteristics for the 1868 patients in the primary aim cohort are shown in Table 1. There were 974 (52%) patients who were not given a reversal agent and 894 (48%) who received a reversal agent.

**Table 1.**
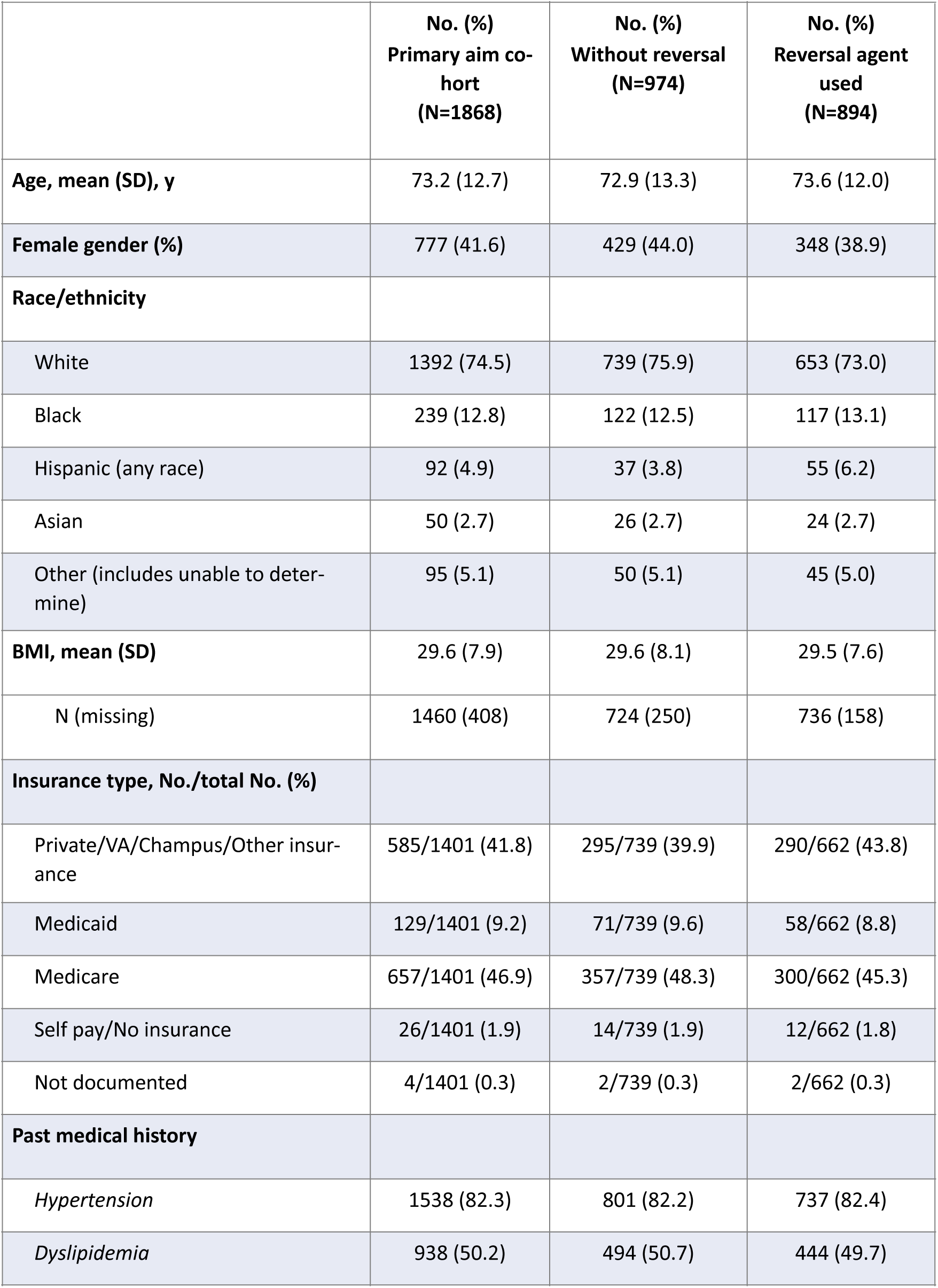
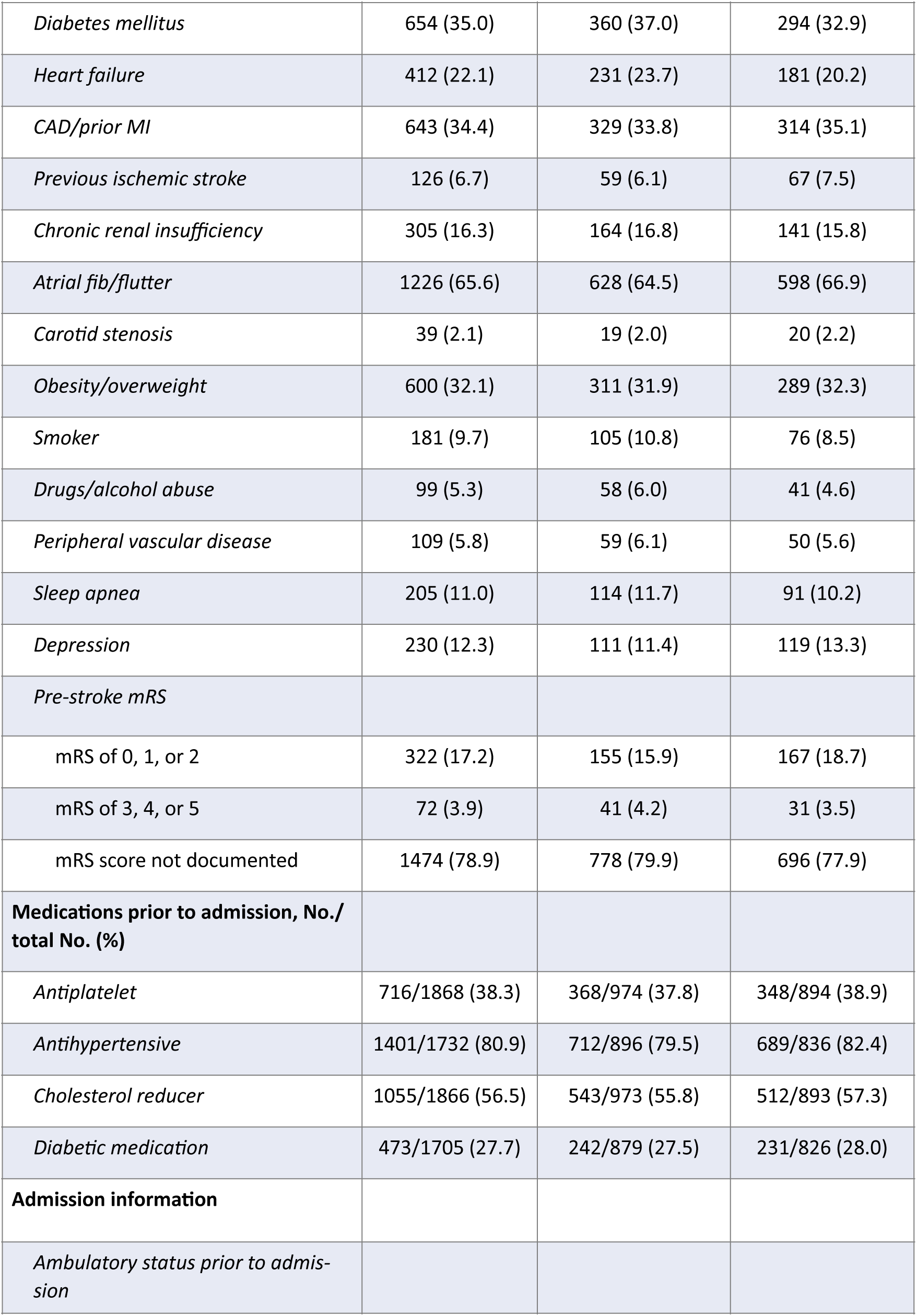
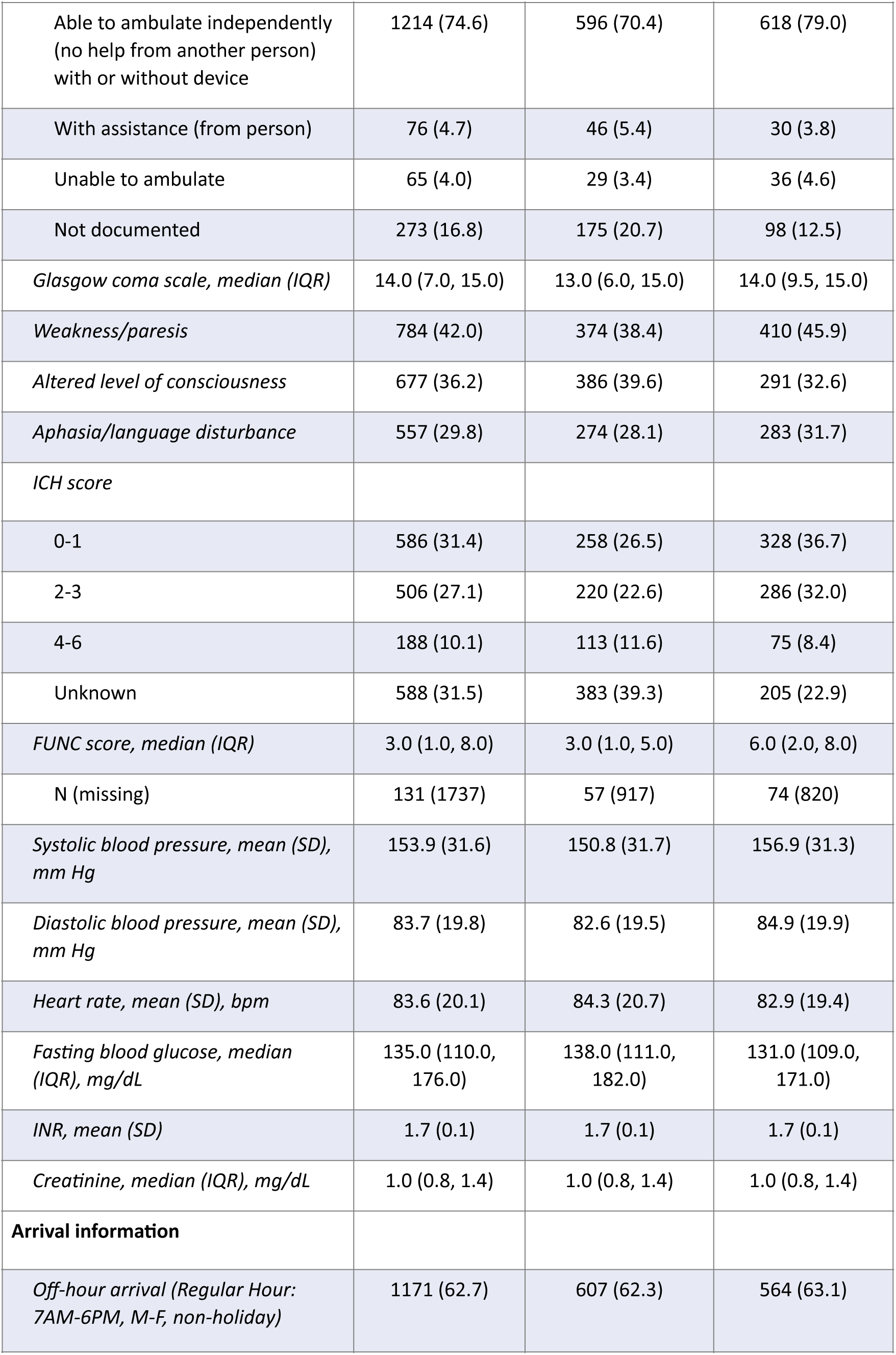
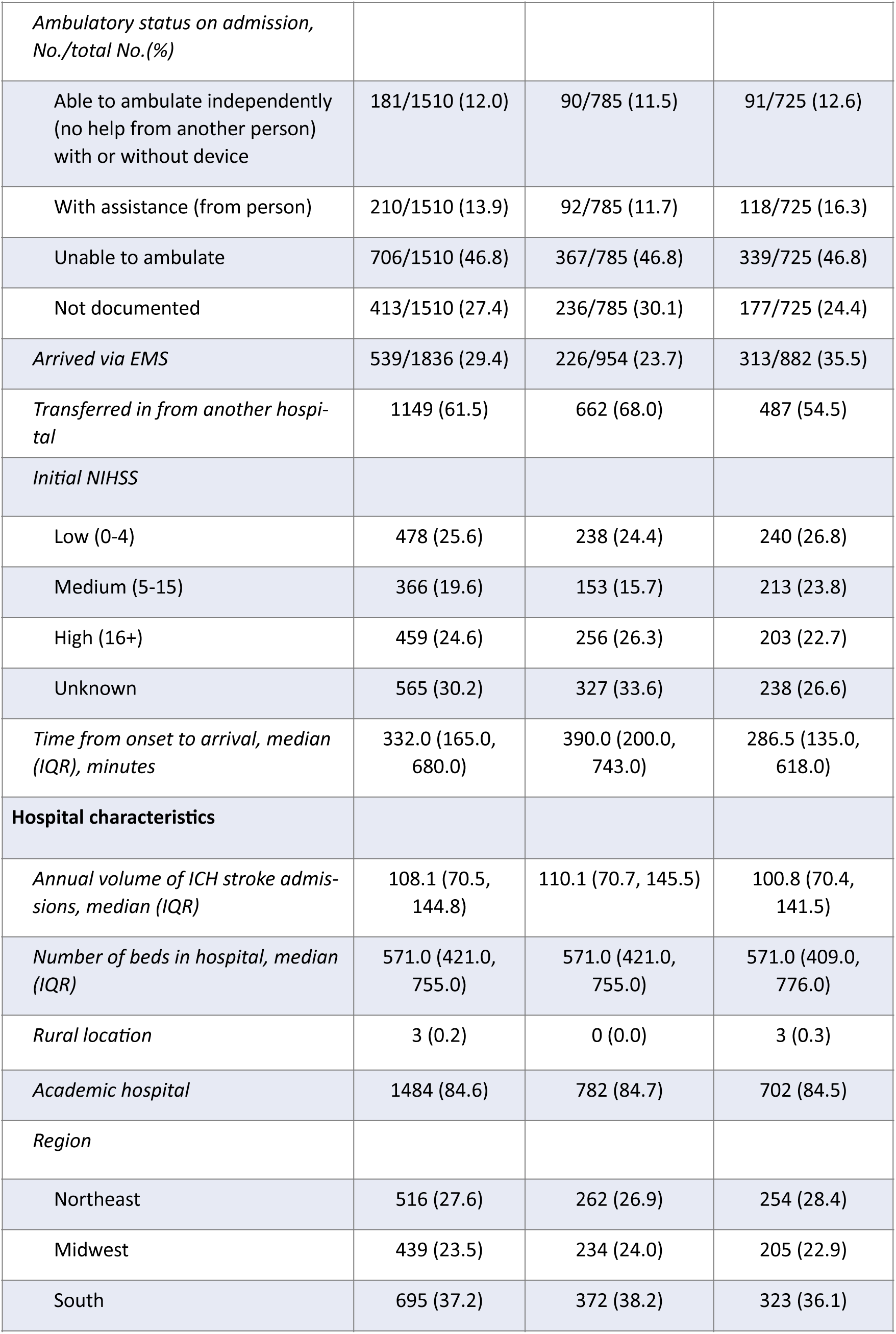
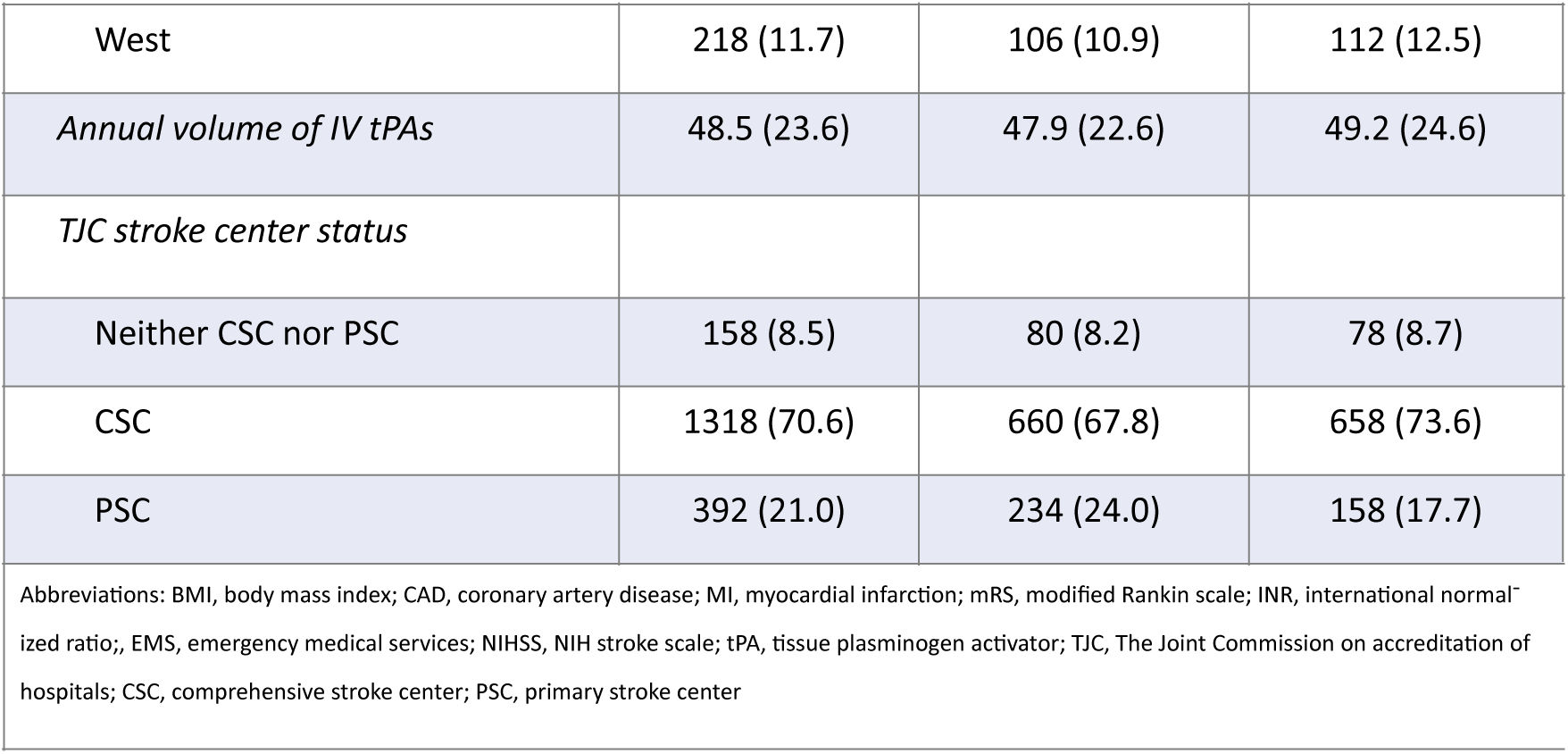
Patient and Hospital Characteristics between those Exposed to a Reversal Agent Versus Not Exposed. – **Primary Aim Cohort.**

The mean age was 73 years, 42% were female, and 75% of patients identified as white. The observed mean BMI was on the threshold of obesity at 29.6 kg/m^2, but 22% of patients were missing BMI (see Supplementary Table 3 for more details on missingness rates). Medicare and private insurance were the most common payors at 47% and 42%, respectively. Just under 10% of patients were on Medicaid. Hypertension was the most common comorbidity (82%), followed by atrial fibrillation/flutter (66%), and then dyslipidemia (50%). As such, 81% of patients were on an antihypertensive and 57% were on a cholesterol reducer prior to admission.

The primary outcome was discharge mRS 0-3 after reversal versus non-reversal of subtherapeutic warfarin anticoagulation for patients with acute intracerebral hemorrhage. A similar proportion of patients at discharge had mRS 0-3 with reversal or without reversal (21.0% versus 23.1% respectively (aOR 0.80 [95% CI 0.63, 1.005])). The sensitivity analysis of INR <= 1.4 after reversal showed similar results for the primary outcome (aOR 0.82 [95% CI 0.63, 1.06]) Patient and hospital characteristics for the 2569 patients in the secondary aims cohort are displayed in Supplementary Table 1 in the supplemental appendix. There were 1351 (53%) patients who were not given a reversal agent and 1218 (47%) that were. Characteristics were similar to the primary aim cohort. Event rates and adjusted effect sizes are reported in Table 2. The distribution of mRS at discharge by reversal agent is shown in Figure 2. When considering mRS as an ordinal variable the model estimated the odds cumulatively of having a given level of mRS or better by combining that level of mRS with all lower mRS values. Receiving a reversal agent was associated with 21% higher odds of having mRS of 0-4 vs. 5-6 at discharge (95% CI = 1.001, 1.48).

**Figure 2.**
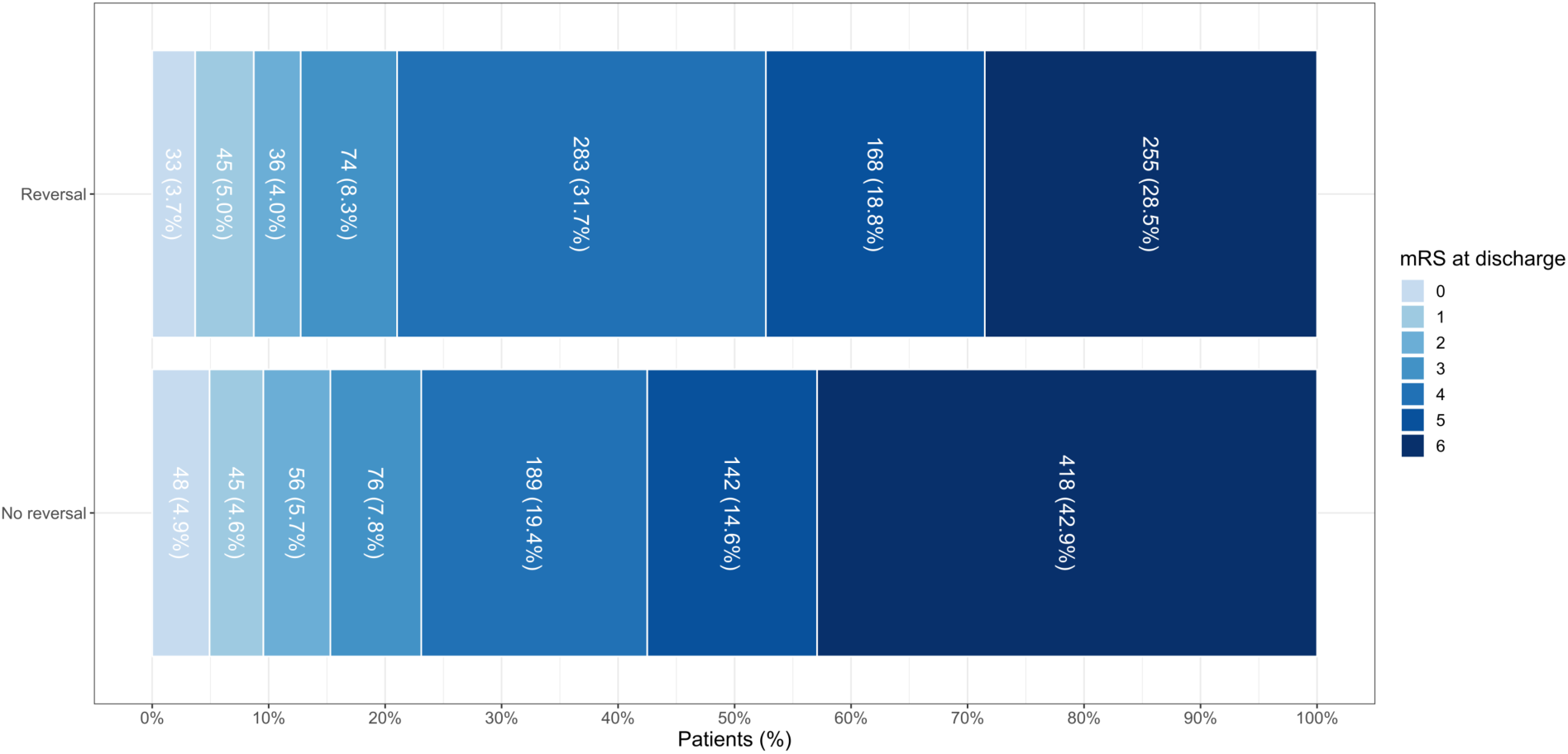
Distribution of mRS at discharge by receipt of a reversal agent. The distribution of mRS at discharge by whether a reversal agent was given is shown in Figure 2. When considering mRS as an ordinal variable we used non-proportional odds cumulative logistic regression model. The model estimated consecutively the odds of having each level of mRS or better by comparing the combination of patients at that level of mRS combined with all lower mRS values. i.e. 0 vs 1-6, 0-1 vs 2-6, etc. Receiving a reversal agent was associated with 21% increased odds of mRS 0-4 vs 5-6 at discharge (95% CI = 1.001, 1.48).

**Table 2.** Association between Reversal Agent Use and Outcomes.

| Table 2. Association between Reversal Agent Use and Outcomes |  |  |  |  |  |
| --- | --- | --- | --- | --- | --- |
|  | No. (%)<br>Overall<br>(N=1868) | No. (%)<br>No reversal<br>agent used<br>(N=974) | No. (%)<br>Reversal<br>agent used<br>(N=894) | Unadjusted<br>effect size<br>(95% confi-<br>dence in-<br>terval) | Adjusted<br>effect size<br>(95% confi-<br>dence in-<br>terval) |
| **Primary** |  |  |  |  |  |
| mRS at discharge (0-3 vs. 4-6) | 413 (22.1) | 225 (23.1) | 188 (21.0) | 0.89 (0.71, 1.10)<sup>1</sup> | 0.80 (0.63, 1.00)<sup>1</sup> |
| **Secondary** |  |  |  |  |  |
| mRS at discharge (ordinal) |  |  |  |  |  |
| 0 | 81 (4.3) | 48 (4.9) | 33 (3.7) | 0.73 (0.48, 1.15)<sup>1</sup> | 0.66 (0.41, 1.07)<sup>1</sup> |
| 1 | 90 (4.8) | 45 (4.6) | 45 (5.0) | 0.90 (0.66, 1.24)<sup>1</sup> | 0.87 (0.62, 1.22)<sup>1</sup> |
| 2 | 92 (4.9) | 56 (5.7) | 36 (4.0) | 0.81 (0.62, 1.05)<sup>1</sup> | 0.77 (0.58, 1.02)<sup>1</sup> |
| 3 | 150 (8.0) | 76 (7.8) | 74 (8.3) | 0.89 (0.71, 1.08)<sup>1</sup> | 0.80 (0.63, 1.00)<sup>1</sup> |
| 4 | 472 (25.3) | 189 (19.4) | 283 (31.7) | 1.51 (1.25, 1.80)<sup>1</sup> | 1.21 (1.00, 1.48)<sup>1</sup> |
| 5-6 | 983 (52.6) | 560 (57.5) | 423 (47.3) | --- | --- |
| In-hospital mortality or discharge to hospice, No./total No. (%) | 930/2563 (36.3) | 558/1346 (41.5) | 372/1217 (30.6) | 0.62 (0.53, 0.73)<sup>1</sup> | 0.75 (0.63, 0.89)<sup>1</sup> |
| Development of DVT or PE, No./total No. (%) | 66/2569 (2.6) | 31/1351 (2.3) | 35/1218 (2.9) | 1.26 (0.77, 2.06)<sup>1</sup> | 1.47 (0.88, 2.46)<sup>1</sup> |
| Length of stay among all patients, median (IQR), days | 5 (3, 10) | 4 (2, 9) | 6 (4, 12) | 1.34 (1.23, 1.46)<sup>2</sup> | 1.25 (1.13, 1.37)<sup>2</sup> |
| Length of stay among those that survived to discharge, median (IQR), days | 6 (4, 12) | 6 (3, 10) | 7 (4, 13) | 1.23 (1.12, 1.34)<sup>2</sup> | 1.17 (1.07, 1.28)<sup>2,3</sup> |
| Home, No./total No. (%) | 516/2563 (20.1) | 292/1346 (21.7) | 224/1217 (18.4) | 0.81 (0.67, 0.99)<sup>1</sup> | 0.79 (0.65, 0.97)<sup>1</sup> |
| Hospice/death, No./total No. (%) | 930/2563 (36.3) | 558/1346 (41.5) | 372/1217 (30.6) | 0.62 (0.53, 0.73)<sup>1</sup> | 0.75 (0.63, 0.89)<sup>1</sup> |
| Skilled nursing, No./total No. (%) | 467/2563 (18.2) | 211/1346 (15.7) | 256/1217 (21.0) | 1.43 (1.17, 1.75)<sup>1</sup> | 1.33 (1.08, 1.65)<sup>1</sup> |
| Rehab, No./total No. (%) | 557/2563 (21.7) | 254/1346 (18.9) | 303/1217 (24.9) | 1.43 (1.18, 1.72)<sup>1</sup> | 1.20 (0.98, 1.47)<sup>1</sup> |
| Fully ambulatory at discharge, No./total No. (%) | 551/1801 (30.6) | 313/877 (35.7) | 238/924 (25.8) | 0.63 (0.51, 0.77)<sup>1</sup> | 0.68 (0.54, 0.85)<sup>1,3</sup> |
| <sup>1</sup> Odds ratio<br><sup>2</sup> Risk ratio<br><sup>3</sup> Model included GCS, ICH score, and NIHSS due to imbalance between treatment arms in sub-group after weighting |  |  |  |  |  |

Patients receiving a reversal agent had 25% lower odds of dying in the hospital or being discharged to hospice (95% CI = 0.63, 0.89) after weighting. LOS was longer for patients that received a reversal agent, both among all patients (median 6 days vs 4 days) and just those that survived to discharge (median 7 days vs 6 days). Receiving reversal was associated with 21% lower odds of being discharged to home (95% CI = 0.65, 0.97) and 33% higher odds of being discharged to skilled nursing (95% CI = 1.08, 1.65). Re^-^ versal was also associated with 32% lower odds of being fully ambulatory at discharge (95% CI = 0.54, 0.85). Patients that received a reversal agent did not have significantly different odds of developing DVT/PE or being discharged to rehabilitation facility.

### Association with ICH Score

Among those with a high (4-6) ICH Score, patients that received a reversal agent had 80% lower odds (95% CI = 0.08, 0.51) of dying in the hospital or being discharged to hospice. There were no unreversed patients with an ICH Score of 4-6 that had a 0-3 mRS at discharge, most of those patients died in the hospital. Similarly, there were no unreversed patients with an ICH score of 4-6 that were fully ambulatory at discharge. Even among those who received a reversal agent and had an ICH Score of 4-6, only 2/28 (7%) were fully ambulatory at discharge.

Among those with a low (0-1) or medium (2-3) ICH Score core reversal was not associated with in-hospital mortality or discharge to hospice after weighting. Among those with an unknown ICH Score, patients that received a reversal agent had lower odds of dying in the hospital or being discharged to hospice (aOR = 0.62; 95% CI = 0.46, 0.85), but it’s difficult to know who these patients are or why they are missing ICH Score.

### Association with withdrawal of life-sustaining treatments

Among the patients who died, a greater proportion of those without reversal were classified as receiving comfort measures only (CMO) 373/418 (89.2%) versus 203/255 (79.6%) where reversal was given.

## Discussion

Reversal of sub-therapeutic warfarin anticoagulation after acute spontaneous intracerebral hemorrhage was not associated with improved short-term functional outcomes based upon an expected predetermined threshold for highly morbid neurologic illness. The primary outcome did not show effect of reversal of subtherapeutic warfarin on the proportion of patients with mRS 3 or lower in patients with acute spontaneous intracerebral hemorrhage. An mRS of 4 represents moderately severe disability, characterized by patients being unable to walk without assistance and unable to attend to their own bodily needs without assistance. It is at mRS 4 that we see an association with subtherapeutic warfarin reversal and outcome. The apparent shift in functional outcome from death to disability is reminiscent of the effect of other treatments in highly morbid neurologic disease, such as decompressive hemicraniectomy for traumatic brain injury, decompression of massive ischemic stroke, and surgical treatment of ICH. See Figure 2.

In this study, fewer patients with warfarin reversal had combined in-hospital mortality or discharge to hospice (30.6% vs 41.5%). Compared to those without reversal, the greatest effect is in highly morbid patients. Patients with a high (4-6) ICH score that received a reversal agent had 80% lower odds (95% CI = 0.08, 0.51) of dying in the hospital or being discharged to hospice. Reversal was associated with increased likelihood of discharge to a nursing facility, decreased likelihood of discharge to home, decreased likelihood of being ambulatory at discharge, and increased length of stay.

The GWTG-Stroke registry does not report data on hematoma expansion so it cannot be determined whether the association between reversal and survival was related to actual lessening of the hematoma expansion. The reversal is believed to have been effective in changing the INR based upon on the sensitivity analysis where there was only 22% crossover of reversed patients to not having corrected INR. A subgroup analysis would be able to determine if the reversed patients who did not achieve corrected INR contributed to the disability or death in that cohort of reversed patients. The specific outcomes of those who failed to be reversed is not known, and the specific timing and type of reversal agent is not known, Because of the observational nature of this study we cannot exclude the possibility that the results may have been affected by residual or unmeasured confounding, even though propensity weighting was used to control for confounding as best as possible. One potential source of confounding could be differential use of limitation of care orders or early withdrawal of care. It is possible that patients who received reversal were more likely to receive future life-sustaining care of all kinds. There were imbalances between the reversed and non-reversed groups that suggest that the reversed and non-reversed patients may have been treated with different intensities, however that evidence is weak. Treating physicians and families may have opted not to treat the more severe cases as aggressively.

The ICH Score is a measure of severity in intracranial hemorrhage; the ICH Score was unknown in 31.5% of patients making any inference from the ICH Scores speculative, but the ICH Scores were higher among the unreversed patients. Similarly, the FUNC score is designed as a predictor of functional recovery after ICH. The FUNC score was worse in the unreversed patients in whom it was known, however the FUNC score was only recorded in 7% of patients.

The time to reversal is an important potential confounder. The duration of time between ictus of the bleed and patient time of arrival is partially corrected for by the inclusion of stroke center type and inclusion of arrival by EMS in the propensity models. The comprehensive stroke centers were more likely to have received referral patients who may not have received reversal before transportation. The time elapsed is greater for patients transferred from another hospital versus patients arriving directly by EMS. To some degree the patients with subtherapeutic INR may represent patients who already have greater time since onset of injury to presentation. This can be seen comparing this study cohort’s subtherapeutic patients with other patients from the registry with normal INR or therapeutic INR. As the INR range decreases there is a progressively lower proportion of patients arriving by EMS [57.3% (INR > 1.9), 29.4% (INR 1.5-1.9), 15.0% (INR <1.5)], and there is a progressively higher proportion of patients transferred in from another hospital as the INR range decreases [29.8% (INR > 1.9), 61.5% (INR 1.5-1.9), 80.2% (INR<1.5)].

With reversal there was an apparent benefit among patients with mRS 4 and 5, and appeared to represent a shift from death to disability. However, mRS at discharge is a short-term outcome, and it may not be reflective of the eventual state of recovery a patient might achieve.^25,26^ Effects on functional outcome may take more time to manifest in the severely injured patients, and it may be too early to tell their eventual state of recovery.

The question addressed in this study is an important one as there appears to be clinical equipoise given the similar proportions of patients who were treated or not treated. While the use of vitamin K antagonists is decreasing over time, they are still encountered frequently in clinical practice. There remain several subgroups of patients in whom vitamin K antagonists are preferred over direct oral anticoagulants, including patients with metallic heart valves, ventricular assist devices, severe renal disease, antiphospholipid antibody syndrome, and gastrointestinal malignancies. Therefore, managing vitamin K antagonist-associated ICH will remain an important part of clinical practice for the foreseeable future. Of the patients who present with vitamin K antagonist-associated ICH, approximately 25% have INR <2.^27^ This analysis provides the clinician with information, albeit not from a controlled trial, on the potential benefits and risks of reversing INR in the subset with INR 1.5-1.9. Given the decreasing overall use of vitamin K antagonists and the lack of novel agents in this class, it seems unlikely that there will be future randomized controls trials to provide definitive data.

Reversal of sub-therapeutic INR appears safe with few thrombotic complications. Reversal is associated with lower mortality. Reversal is associated with survival at higher disability mRS than was expected. There are several intriguing effects seen in this data: the mortality benefit, the lack of short-term functional outcome benefit at the expected level, more patients with higher degree of functional disability at the time of discharge, fewer patients walking at discharge, and longer hospital stays. Functional improvement over time is expected to occur but is not known from this study. The functional outcome at discharge is not long enough to determine full potential for recovery. Satisfaction scores for the patients and families long-term are unknown.

### Limitations

This study utilizes data that was abstracted for non-research purposes and analysis is only possible for the existing data. The mRS at the time of discharge is a meaningful, yet imperfect, outcome.^25^ An outcome measure at fuller recovery would be preferable but harder to obtain without greater loss to follow-up. An alternative would be to use projections of long-term outcomes derived from a smaller data set of 6-month recovery.^35^

The outcomes may be affected by uncontrolled effects of physicians’ decision to treat and family decisions regarding withdrawal of live-sustaining treatment. However without specific information regarding end-of-life decision making, any inference about decision-making is tentative.

The final determination of the meaningfulness of an outcome is best interpreted by the patients and families themselves. Thus, the absence of patient reported outcomes remains a short-coming.

### Conclusion

Reversal of subtherapeutic warfarin anticoagulation in patients with acute intracerebral hemorrhage and INR 1.5 - 1.9 was not associated with an improvement in short term functional outcome. However, patients who received a reversal agent had 25% lower odds of dying in the hospital or being discharged to hospice, but had a longer hospital stay and were less likely to be fully ambulatory at discharge. Reversal was not associated with an increased risk of thromboembolism. A randomized clinical trial would be needed to further evaluate the potential benefits and risk of reversal in this patient population.

## Sources of funding

“The Get With the Guidelines®—Stroke (GWTG) program is provide by the American Heart Association/American Stroke Association. GWTG—Stroke is sponsored, in part, by Novartis, Novo Nordisk, AstraZeneca, Bayer, Tylenol, Alexion, and AstraZeneca Rare Disease.”

## Disclosures

None.

## Data Availability

The GWTG-Stroke database has independent policies for use of data.

## Abbreviations

aOR: adjusted odds ratio
aRR: adjusted risk ratio
BMI: body mass index
BP: blood pressure
CI: confidence interval
CMO: comfort measures only
DOAC: direct oral anticoagulant
DVT: deep venous thrombosis
EMS: emergency medical services
FUNC: Functional outcome in patients with primary intracerebral hemorrhage score
GCS: Glascow coma scale
GWTG: Get With The Guidelines
GWTG-Stroke: Get With The Guidelines – Stroke
ICH: intracerebral hemorrhage
INR: international normalized ratio
LOC: level of consciousness
LOS: length of stay
mRS: modified Rankin scale
NIHSS: NIH stroke scale
OR: odds ratio
PE: pulmonary embolus
SD: standard deviation

## Notes

### Competing Interest Statement

The authors have declared no competing interest.

### Author Declarations

DCRI IRB approval

